# Predicting Post-Liver Transplant Outcomes in Patients with Acute-on-Chronic Liver Failure using Expert-Augmented Machine Learning

**DOI:** 10.1101/2023.03.03.23286729

**Authors:** Jin Ge, Jean C. Digitale, Cynthia Fenton, Charles E. McCulloch, Jennifer C. Lai, Mark J. Pletcher, Efstathios D. Gennatas

## Abstract

**Background:** Liver transplantation (LT) is a treatment for acute-on-chronic liver failure (ACLF) but up to 40% mortality post-LT has been reported. Existing post-LT models in ACLF have been limited by small samples. In this study, we developed a novel Expert-Augmented Machine Learning (EAML) model to predict post-LT outcomes.

**Methods:** We identified ACLF patients in the University of California Health Data Warehouse (UCHDW). We used EAML, which uses the RuleFit machine learning (ML) algorithm to extract rules from decision-trees that are then evaluated by human experts, to predict post-LT outcomes. We compared EAML/RuleFit’s performances versus other popular models.

**Results:** We identified 1,384 ACLF patients. For death at one-year: areas-under-the-receiver-operating characteristic curve (AUROCs) were 0.707 (Confidence Interval [CI] 0.625-0.793) for EAML and 0.719 (CI 0.640-0.800) for RuleFit. For death at 90-days: AUROCs were 0.678 (CI 0.581-0.776) for EAML and 0.707 (CI 0.615-0.800) for RuleFit. In pairwise comparisons, EAML/RuleFit models outperformed cross-sectional models. Divergences between experts and ML in rankings revealed biases and artifacts in the underlying data.

**Conclusions:** EAML/RuleFit outperformed cross-sectional models. Significant discrepancies between experts and ML occurred in rankings of biomarkers used in clinical practice. EAML may serve as a method for ML-guided hypothesis generation in further ACLF research.

## Introduction

Acute-on-chronic liver failure (ACLF) is commonly defined as acute decompensation of end-stage liver disease (ESLD) with extra-hepatic organ failure and is associated with high short-term mortality.(2–7) Liver transplantation (LT) is a well-established treatment for patients with ACLF who are refractory to supportive care and treatment for the underlying precipitant. Due to critical illness, however, LT is estimated to be feasible in only 25% of ACLF patients.(8) Moreover, there have been conflicting post-LT outcomes reported for ACLF patients with some sub-populations having up 40% three-month mortality.(9,10) There is an unmet need for tools to predict post-LT outcomes for ACLF patients in the pre-LT setting (and without intra-operative or post-LT data) to ensure utility.(11,12)

Multiple international research consortia, such as the North American Consortium for the Study of End-Stage Liver Disease (NACSELD),(3) the European Association for the Study of the Liver-Chronic Liver Failure Consortium (EF-CLIF),(4) and the Asian Pacific Association for the Study of the Liver ACLF Research Consortium (APASL ACLF);(13) have developed scoring systems to predict pre-LT outcomes. None of these models, however, specifically evaluates for post-LT outcomes. One of the few models that specifically evaluate for post-LT outcomes is the Transplantation for ACLF-3 Model (TAM) score, which was trained on a cohort of 76 patients with EF-CLIF grade-3 (severe) ACLF at a single French center and validated in 76 patients in four other centers.(14) Despite its potential utility, the TAM model has not been studied in non-European settings outside of limited cohorts.

In addition, one of the major barriers to building post-LT outcomes models for ACLF patients is that ACLF is a heterogeneous and dynamic clinical syndrome, as evidenced by diverging definitions in different geographies.(3,4,6,8,13) Existing prediction models do not utilize vast numbers of data features available in electronic health records (EHRs) to better define dynamic clinical trajectories seen in patients with ACLF. Our group had previously demonstrated an informatics approach to extract EHR data that yielded a median of 454 features per admission to more accurately represent ACLF patients’ clinical courses.(15) Machine Learning (ML) is well-suited for analyzing such data, but can be misleading when taken out of context of biological or clinical mechanisms.(16,17)

Expert-Augmented Machine Learning (EAML) is an emerging technique that overcomes this limitation of ML by extracting rules from decision-tree ML models for human expert feedback. EAML has two potential benefits: 1) To create combined models that incorporate the best of human and ML knowledge, and 2) To evaluate for differences between humans and ML. These differences could represent human biases (e.g., experts ignoring important variables identified by ML) or artifacts in the underlying data (e.g., experts are identifying the important variables but there is over-representation of other clinical characteristics in this population not seen elsewhere).

In this study, we utilized a novel multi-center EHR database, the University of California Health Data Warehouse (UCHDW), to construct an EAML model to predict post-LT outcomes in patients with ACLF.

## Methods

### The University of California Health Data Warehouse (UCHDW)

The UCHDW is a unique data asset created from the EHRs and claims data from the five major University of California Health (UCH) Medical Centers (Davis, Irvine, Los Angeles, San Diego, and San Francisco) and managed by the Center for Data-Driven Insights and Innovation (CDI2).(1) UCHDW holds data on 6.2+ million well-characterized patients seen at UCH since 2012. All data in UCHDW are harmonized in the Observational Medical Outcomes Partnership (OMOP) common data model, version 5.3.1.(18) All data elements in UCHDW are de-identified prior to the receipt by end-users with no clinical notes or imaging. UCHDW has previously been utilized to analyze treatment utilization patterns between UCH health systems and amongst individual providers within each health system.(19) For all analyses, we utilized UCHDW, versioned as of September 22, 2022 and accessed on October 20, 2022.

### Study Population

We isolated all adults (>= 18 years) who underwent an orthotopic liver transplantation procedure, as defined by the OMOP concept identifiers 2109321 (CPT4 code) or 4067458 (SNOMED code), based on the ATHENA OMOP vocabulary dictionary,(20) in UCHDW between January 1, 2013 through December 31, 2021. We included patients who underwent multi-organ (such as simultaneous liver-kidney transplant) and re-transplant procedures as they may have been in ACLF prior to transplant. Consistent with prior informatics approaches for detecting ACLF admissions, we excluded all patients who underwent transplant within 48 hours of admission as they were likely admitted electively.(15) We included patients who had evidence of ACLF prior to the time of LT through a previously published informatics-driven approach.(15) Briefly, this involves identifying any patient who meets ACLF diagnostic criteria based on the NACSELD or EF-CLIF definitions prior to LT. We did not use the APASL ACLF diagnostic criteria due to bacterial infection being the most common precipitant of ACLF in patients with cirrhosis in the United States.(21,22)

### Measurements

We extracted all structured clinical information associated with the admission of interest. Baseline characteristics included age, sex, race/ethnicity, height, weight, body mass index, and censored identity of the UCH facility (defined as “UC-1,” “UC-2,” and “UC-3”). Laboratory measurements, liver disease etiologies, complications of cirrhosis, comorbid medical conditions, dialysis state, ventilation parameters, and vasopressor administration were extracted based on previously defined OMOP concept identifiers.(20,23)

As patients may have different lengths of stay before LT, we focused only on data values from the day of admission and the day before LT. We dropped measurements from other time points from consideration. Continuous data features were averaged by hospitalization day. We defined changes between admission and transplant based on the differences between data features between admission and day before LT.(24–26) All intra-operative data values and values after transplant were excluded from our analyses as our intent was to develop a predictive model utilizing only pre-transplant data. Missing data features and variables underwent single imputation with chained random forests, which has been shown to produce low errors and good performances in previous studies utilizing EHR data.(27–29)

### Outcomes

The primary outcome was all-cause mortality at one-year after LT. The secondary outcomes included: 1. All-cause readmissions within 90-days, and 2. All-cause mortality within 90-days after LT. Death was ascertained based on synchronized data with the California Death Registry.(1)

### Model Development and Expert-Augmented Machine Learning (EAML)

The sample of ACLF patients isolated from UCHDW was split by random sampling into training, validation, and test sets in a 60:20:20 ratio.(30–32) The training set was used to fit the model, the validation set was utilized to tune hyperparameters, and the test set was held-out for independent testing. RuleFit training and testing plots are shown in Supplemental Figures 1, 2, and 3 for our three outcomes. We then utilized EAML, as implemented in the rtemis R package, version 0.91, to train one ML model for each of our primary and secondary outcomes of interest (total of three models).(33) rtemis is a platform for advanced ML research and applications, which incorporates several algorithms, including EAML.(34)

**Figure 1.**
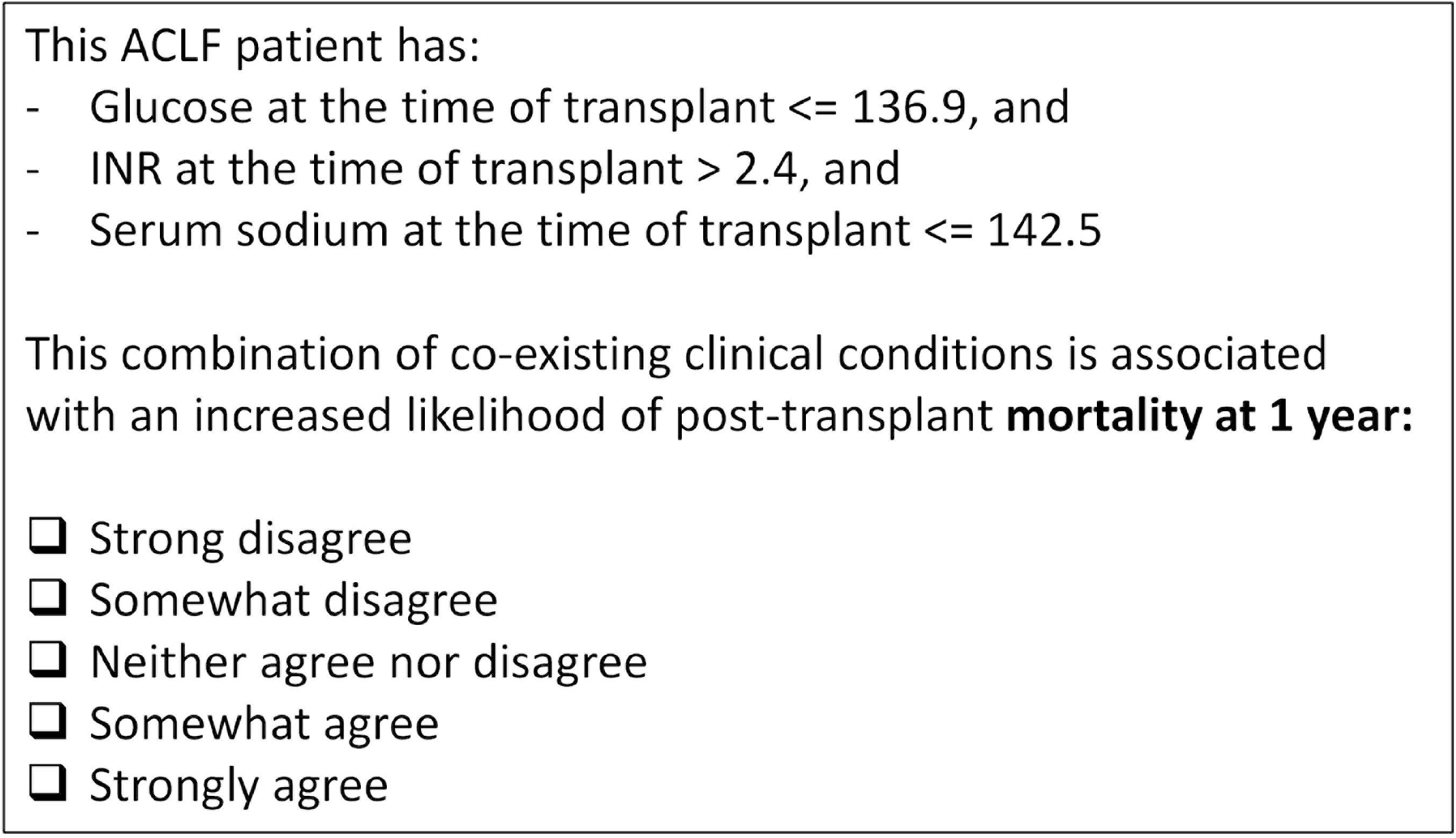
Example Survey Question Utilized to Obtain Expert Input

**Figure 2.**
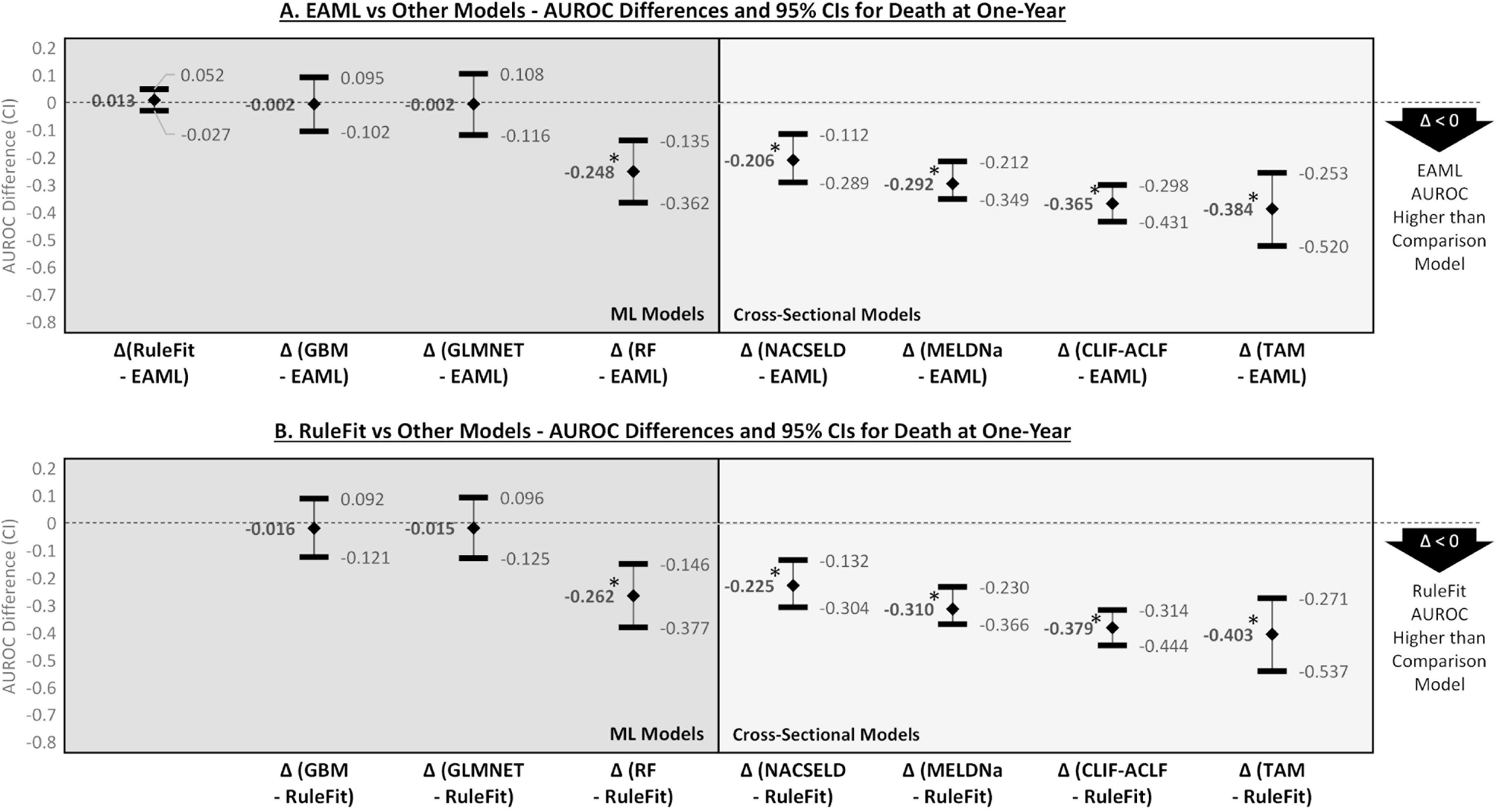
AUROC Differences and Confidence Intervals for EAML/RuleFit versus Other Models for the Outcome of Death at One-Year

**Figure 3.**
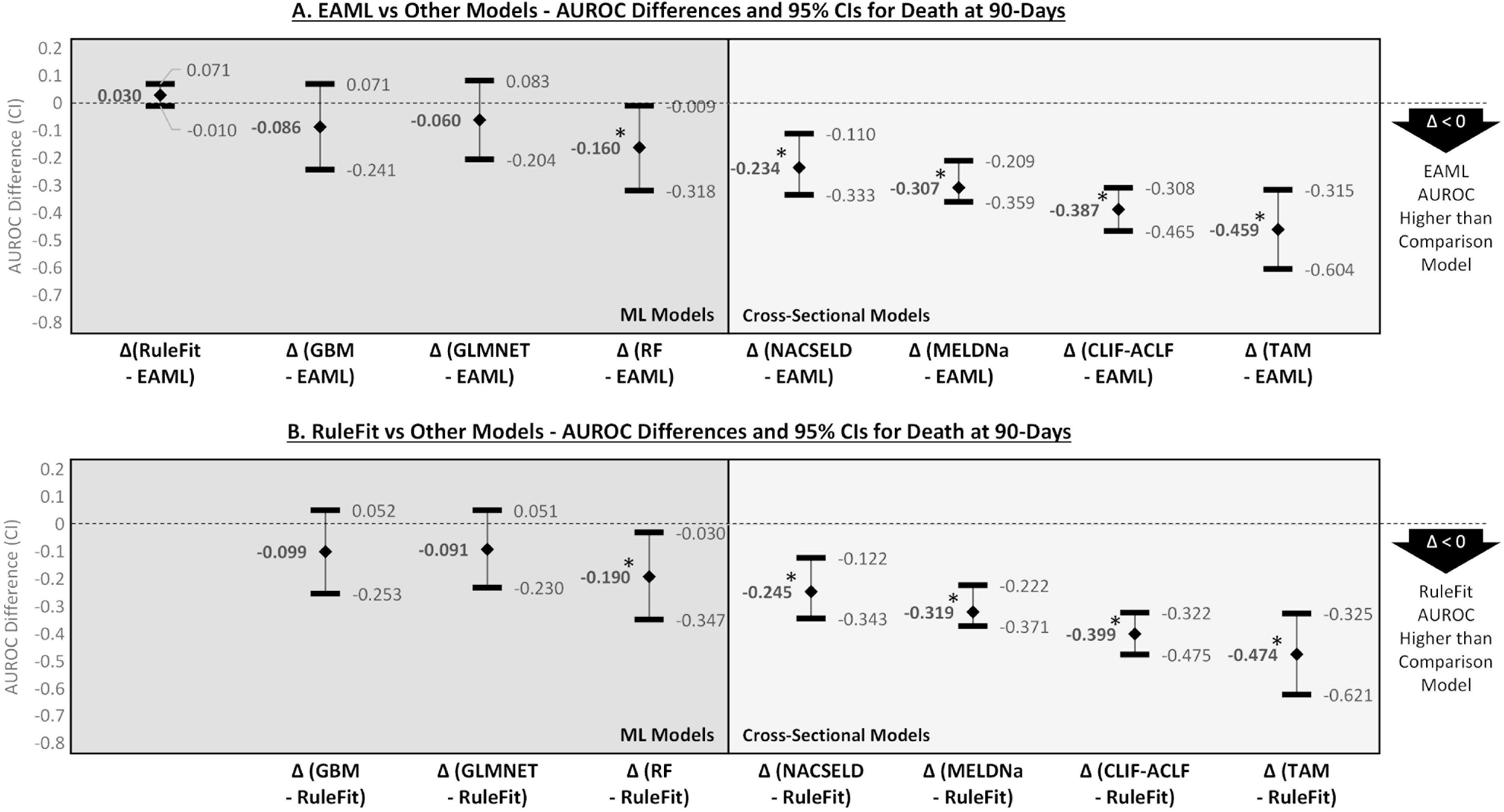
AUROC Differences and Confidence Intervals for EAML/RuleFit versus Other Models for the Outcome of Death at 90-Days

As described above, EAML is an ensemble ML algorithm that incorporates human knowledge by converting high-dimensional training data into Likert-scale questions.(33) EAML first trains a predictive model using the RuleFit algorithm,(35) which is a combination of a Gradient Boosting Machine (GBM) decision-tree model (trained on the data to generate rules), and a Least Absolute Shrinkage and Selection Operator (LASSO) model (used to select rules generated by the GBM model).(35) The RuleFit model training outputs include the detailed rules, model coefficients (represents the change in response associated with the rule), and empirical risk (rating of the rule importance by the machine).

Utilizing the rules selected by RuleFit, we then created an online survey on the Qualtrics platform (example question in Figure 1) that was sent to 15 hepatologists throughout the world who conduct clinical care and research in ACLF recruited from a convenience sample. These experts were asked to rate rules on a 5-point Likert-scale based on perceived associations with the outcomes of interest. We calculated expert rankings based on the averages of these ratings. We then took the differences in rankings between the experts and those generated by the RuleFit model to calculate penalties. These penalties were then incorporated into the RuleFit models by eliminating the top quartile of the most discrepant rules (highest fourths of absolute rank differences between RuleFit and expert rankings) to create the EAML models for each of the three outcomes.(33)

### Statistical Analyses and Model Performance Evaluation

Clinical characteristics and laboratory data were summarized by medians and interquartile ranges (IQR) for continuous variables or numbers and percentages (%) for categorical variables. Comparisons between the training, validation, and test sets were performed using chi-square and Kruskal-Wallis tests where appropriate.

We evaluated the performances of EAML (with expert input) and RuleFit (without expert input) models through area-under-the-receiver-operating characteristic curve (AUROC), which has been used previously to evaluate ML models in transplant hepatology.(36–39) To compare the performances of the EAML and RuleFit models versus cross-sectional models (MELDNa, NACELD-ACLF, CLIF-C-ACLF, TAM) and other ML algorithms (Random Forest [RF], GBM, and Elastic-Net Regularized Generalized Linear Model [GLMNET]), we calculated AUROC differences between each pair of models (e.g., AUROC differences between EAML and NACSELD) and their confidence intervals using bootstrapping with 2,000 iterations per pair-wise comparison.(40,41) We calculated MELDNa, NACSELD-ACLF, CLIF-C-ACLF, and TAM scores per previously published literature.(3,4,14,42) We used rtemis implementations of RF, GBM, and GLMNET to generate comparison ML models.

All data queries, extractions, and transformations of OMOP concept identifiers in UCHDW were conducted using the Microsoft Azure implementations of Spark, version 2.12. All statistical analyses were performed utilizing Spark-R, version 4.1.3 “One Push-Up” (R Core Team, Vienna, Austria), and R packages previously noted and documented in the supplemental materials.(43) Two-sided p-values <0.05 were considered statistically significant in all analyses. The use of UCHDW data for this study was authorized by the Institutional Review Board at the University of California, San Francisco under #20-32717 for model generation and #22-37555 for expert input.

## Results

A total of 1,384 patients with ACLF were identified from UCHDW from January 1, 2013 through December 31, 2021. Of the 1,384: 611 (44.1%) were women, 576 (41.6%) Hispanic, 472 (34.1%) non-Hispanic White, 138 (10.0%) Asian, 60 (4.3%) Black, and 122 (8.8%) of Unknown/Other race/ethnicity. Distribution of patients by University of California sites were 410 (29.6%) at UC-1, 173 (12.5%) at UC-2, and 801 (57.9%) at UC-3.

The patients were randomly divided based on a 60:20:20 ratio with 841 patients in the training set, 255 in the validation set, and 288 in the test set. The three sets were broadly similar across multiple demographic and clinical characteristics (e.g. age, race/ethnicity, liver disease etiologies, comorbid conditions, and distribution between UCH facilities). Of note, the median MELDNa scores at admission were 34 (interquartile range [IQR] 29-39), 34 (IQR 30-38), and 34 (IQR 30-38) for the training, validation, and test sets, respectively. Detailed patient characteristics at time of admission are reported in Table 1.

**Table 1.** Baseline Clinical and Demographic Characteristics of the Training, Validation, and Test Set Populations.

|  | <u>Train (N = 841)</u> | <u>Validation (N = 255)</u> | <u>Test (N = 288)</u> | <u>p-Value</u> |
| --- | --- | --- | --- | --- |
| **Female** | 370 (44) | 106 (42) | 135 (47) | 0.46 |
| **Age (IQR)** | 57.5 (49.1-63.8) | 56.2 (46.2-62.9) | 58.0 (47.4-64.5) | 0.19 |
| **UC Health Site** |  |  |  | 0.64 |
| UC-1 | 251 (30) | 70 (27) | 89 (31) |  |
| UC-2 | 110 (13) | 34 (13) | 29 (10) |  |
| UC-3 | 480 (57) | 151 (59) | 170 (59) |  |
| **Race/Ethnicity** |  |  |  | 0.21 |
| Hispanic | 357 (42) | 107 (42) | 112 (39) |  |
| White | 284 (34) | 80 (31) | 108 (38) |  |
| Asian | 89 (11) | 19 (7) | 30 (10) |  |
| Black | 34 (4) | 13 (5) | 13 (5) |  |
| Unknown/Other | 68 (8) | 33 (13) | 21 (7) |  |
| **Etiology of Liver Disease** |  |  |  |  |
| Alcohol Associated | 293 (35) | 90 (35) | 94 (33) | 0.76 |
| Nonalcoholic fatty | 207 (25) | 63 (25) | 73 (25) | 0.97 |
| Hepatitis C | 207 (25) | 61 (24) | 66 (23) | 0.84 |
| Hepatitis B | 87 (10) | 17 (7) | 24 (8) | 0.17 |
| Autoimmune | 67 (8) | 24 (9) | 23 (8) | 0.75 |
| **Previous Complications of Cirrhosis** |  |  |  |  |
| Ascites | 746 (89) | 231 (91) | 258 (90) | 0.68 |
| Hepatic Encephalopathy | 636 (76) | 191 (75) | 214 (74) | 0.90 |
| Esophageal Varices | 468 (56) | 139 (55) | 170 (59) | 0.51 |
| Spontaneous Bacterial Peritonitis | 179 (21) | 60 (24) | 70 (24) | 0.50 |
| Hepatocellular Carcinoma | 113 (13) | 35 (14) | 46 (16) | 0.56 |
| **Comorbidities** |  |  |  |  |
| Chronic Renal Failure | 476 (57) | 131 (51) | 173 (60) | 0.12 |
| Diabetes | 362 (43) | 107 (42) | 120 (42) | 0.90 |
| Coronary Artery Disease | 231 (27) | 60 (24) | 84 (29) | 0.31 |
| Congestive Heart Failure | 130 (15) | 32 (13) | 41 (14) | 0.50 |
| **Laboratory Tests** |  |  |  |  |
| **MELDNa** | 34.1 (29.0-39.1) | 33.8 (30.1-38.1) | 33.7 (29.5-37.6) | 0.90 |
| Sodium | 134.0 (129.0-138.0) | 135.0 (130.0-139.0) | 134.0 (129.0-138.0) | 0.24 |
| Creatinine | 2.0 (1.2-3.4) | 2.0 (1.2-3.1) | 2.0 (1.3-3.2) | 0.74 |
| Albumin | 3.1 (2.6-3.7) | 3.1 (2.6-3.6) | 3.2 (2.6-3.6) | 0.50 |
| Aspartate Transferase | 70.0 (43.5-125.0) | 37.0 (23.0-72.0) | 36.0 (21.0-62.0) | 0.30 |
| Alanine Transferase | 36.0 (21.0-66.5) | 37.0 (23.0-72.0) | 36.0 (21.0-62.0) | 0.76 |
| Alkaline Phosphatase | 111.0 (78.8-163.0) | 117.5 (82.0-177.8) | 110.0 (79.0-156.0) | 0.17 |
| Total Bilirubin | 12.0 (4.6-24.5) | 11.6 (5.2-19.8) | 11.3 (4.3-22.0) | 0.51 |
| White Blood Cell Count | 7.2 (5.1-11.3) | 7.7 (5.0-12.1) | 7.5 (5.0-11.3) | 0.75 |
| Hemoglobin | 8.7 (7.8-10.2) | 8.8 (7.9-10.0) | 8.8 (7.8-10.1) | 0.87 |
| Platelet | 53.0 (37.0-88.5) | 54.0 (37.0-87.3) | 53.0 (35.0-79.0) | 0.41 |
| International Normalized Ratio | 2.3 (1.8-2.9) | 2.3 (1.9-3.1) | 2.3 (1.8-3.0) | 0.35 |
| **Infection** | 110 (13) | 36 (14) | 46 (16) | 0.47 |
| **Hemodialysis** | 60 (7) | 17 (7) | 29 (10) | 0.22 |
| **NACSELD-ACLF** | 559 (66) | 160 (63) | 183 (64) | 0.44 |
| **CLIF-ACLF** |  |  |  | 0.34 |
| Grades 1-2 | 178 (21) | 65 (26) | 64 (22) |  |
| Grade 3 | 663 (79) | 190 (75) | 222 (77) |  |
| **TAM (CLIF-ACLF 3 Only)** |  |  |  | 0.69 |
| 0-1 | 113 (17) | 31 (16) | 39 (18) |  |
| 2 | 120 (18) | 27 (14) | 28 (13) |  |
| 3 | 37 (6) | 10 (5) | 12 (5) |  |

### Primary and Secondary Outcomes

In the total sample of 1,384 patients: 149 (10.8%) met the primary outcome of death at one-year, 97 (7%) met the secondary outcome of death at 90-days, and 621 (44.9%) met the secondary outcome of readmission within 90-days. Distributions and prevalence of the primary and secondary outcomes were similar between the training, validation, and test sets; and are reported in Table 2.

**Table 2.** Outcomes of the Training, Validation, and Test Set Populations.

|  | <u>Train (N = 841)</u> | <u>Validation (N = 255)</u> | <u>Test (N = 288)</u> | <u>p-Value</u> |
| --- | --- | --- | --- | --- |
| **Outcomes** |  |  |  |  |
| Death at one-year | 87 (10) | 28 (11) | 34 (13) | 0.78 |
| Death at 90-days | 55 (7) | 19 (7) | 23 (9) | 0.68 |
| Readmission at 90-days | 367 (44) | 127 (50) | 127 (50) | 0.21 |

### RuleFit and Expert Augmentation

After identification and division of the ACLF patient population as above, we then applied the RuleFit algorithm. RuleFit generated 20 rules for the primary outcome of death at one-year (Table 4), 18 rules for the secondary outcome of death within 90-days (Table 5), and 6 rules for the secondary outcome of readmission within 90-days (Table 6). The rules generated by RuleFit for each of the outcomes were then distributed to 15 hepatologists throughout the world who conduct clinical care and research in ACLF who rated the importance of rules based on a 5-point Likert scale. The aggregated physician rankings along with rank differences between RuleFit and experts are also reported in Tables 4, 5, and 6 for each of the three outcomes. Of note, the greatest discrepancies between RuleFit and human experts occurred in the rankings of biomarkers more commonly utilized in clinical practice, such as age and MELDNa score.

**Table 3.** RuleFit and Expert Rankings for the Primary Outcome of Mortality at One-Year.

| Rule # | Rule Description | # Cases | Model Coefficient | Empirical Risk (Importance) | RuleFit Importance | Expert Importance | Rank Difference |
| --- | --- | --- | --- | --- | --- | --- | --- |
| 1 | Serum Glucose(1) <= 136.92 AND INR(1) > 2.41 AND Serum Sodium(1) <= 142.50 | 229 | 0.80 | 0.97 | 1 | 17 | -16 |
| 2 | Serum Glucose(1) <= 136.92 AND Serum Sodium(1) > 142.50 | 37 | -0.38 | 0.76 | 18 | 20 | -2 |
| 3 | Serum Glucose(1) > 136.92 | 410 | -0.03 | 0.86 | 11 | 19 | -8 |
| 4 | Differences in Serum Albumin <= -0.55 | 84 | 0.72 | 0.96 | 2 | 9 | -7 |
| 5 | Total Bilirubin(1) <= 26.41 AND Differences in Serum Albumin > -0.55 AND Differences in Serum Phosphorus <= 0.98 | 435 | 0.36 | 0.92 | 8 | 10 | -2 |
| 6 | Differences in Serum Albumin > -0.55 AND Differences in Serum Phosphorus > 0.98 | 140 | -0.15 | 0.83 | 14 | 16 | -2 |
| 7 | Serum Sodium(0) <= 138.50 AND Oxygen Saturation(1) <= 98.29 | 232 | 0.62 | 0.96 | 4 | 12 | -8 |
| 8 | Oxygen Saturation(1) > 98.29 AND Differences in Lactate Dehydrogenase > 6 | 135 | -0.61 | 0.79 | 17 | 8 | 9 |
| 9 | Serum Calcium(1) <= 8.45 AND Ionized Calcium(1) <= 1.36 AND Differences in Temperature <= 0.15 | 81 | 0.44 | 0.96 | 3 | 7 | -4 |
| 10 | Ionized Calcium(1) <= 1.36 AND Differences in Temperature > 0.15 | 388 | 0.02 | 0.92 | 9 | 11 | -2 |
| 11 | Ionized Calcium(1) > 1.36 | 77 | -0.62 | 0.79 | 16 | 14 | 2 |
| 12 | Age <= 51.79 | 263 | 0.45 | 0.93 | 7 | 18 | -11 |
| 13 | Alkaline Phosphatase(0) <= 289 AND Differences in Serum Potassium > 1 AND Age > 51.79 | 25 | 0.63 | 0.96 | 5 | 5 | 0 |
| 14 | Alkaline Phosphatase(0) > 289 AND Age > 51.79 | 16 | -0.44 | 0.75 | 19 | 2 | 17 |
| 15 | Alkaline Phosphatase(0) > 63 AND Serum Bicarbonate(0) <= 26.90 AND Differences in Hemoglobin > -0.10 | 283 | -0.35 | 0.85 | 13 | 6 | 7 |
| 16 | Ionized Calcium(0) <= 0.98 AND WBC(1) > 11.89 | 12 | -1.02 | 0.67 | 20 | 3 | 17 |
| 17 | Temperature(0) > 98.05 AND MELDNa(1) <= 32.47 AND Differences in WBC <= 3.90 | 199 | -0.21 | 0.85 | 12 | 4 | 8 |
| 18 | MELDNa(1) > 32.47 | 436 | 0.00 | 0.92 | 10 | 1 | 9 |
| 19 | Serum Glucose(1) > 168 AND Differences in Serum Glucose <= 68.32 AND No New Bacterial Infection (Transplant versus Admission) | 83 | 0.41 | 0.95 | 6 | 15 | -9 |
| 20 | Differences in Serum Glucose > 68.32 | 138 | -0.16 | 0.83 | 15 | 13 | 2 |

(0) Indicates value on the day of admission (1) Indicates value on the day prior to transplant Differences are defined between values on the day prior to transplant versus those on the day of admission

**Table 4.** RuleFit and Expert Rankings for the Secondary Outcome of Mortality at 90-Days.

| Rule # | Rule Description | # Cases | Model Coefficient | Empirical Risk (Importance) | RuleFit Importance | Expert Importance | Rank Difference |
| --- | --- | --- | --- | --- | --- | --- | --- |
| 1 | Blood Urea Nitrogen(1) > 25.50 AND Temperature(1) <= 98.85 AND Differences in Lactate Dehydrogenase <= 40.05 | 324 | 0.39 | 0.97 | 7 | 6 | 1 |
| 2 | Temperature(1) > 98.85 AND Differences in Lactate Dehydrogenase <= 40.05 | 192 | 0.39 | 0.97 | 5 | 5 | 0 |
| 3 | Differences in Lactate Dehydrogenase > 40.05 | 86 | -0.06 | 0.79 | 18 | 4 | 14 |
| 4 | Platelet(1) <= 37.50 AND Temperature(1) <= 100.17 AND Differences in Total Bilirubin <= 5.73 | 232 | 0.72 | 0.98 | 1 | 3 | -2 |
| 5 | Temperature(1) > 100.17 AND Differences in Total Bilirubin <= 5.73 | 24 | -0.10 | 0.79 | 17 | 2 | 15 |
| 6 | Difference in Total Bilirubin > 5.73 | 218 | -0.11 | 0.89 | 12 | 1 | 11 |
| 7 | Serum Calcium(1) > 8.60 AND Hemoglobin(1) > 8.85 AND Differences in MELDNa <= 0.20 | 118 | -0.70 | 0.84 | 16 | 16 | 0 |
| 8 | Heart Rate(1) <= 79.30 | 278 | 0.17 | 0.97 | 4 | 16 | -12 |
| 9 | Serum AST(1) <= 34 AND Heart Rate(1) > 79.30 | 61 | -0.36 | 0.85 | 14 | 13 | 1 |
| 10 | Serum AST(1) > 34 AND Heart Rate(1) > 79.30 AND Differences in Serum Creatinine <= -1.95 | 94 | -0.26 | 0.84 | 15 | 13 | 2 |
| 11 | Serum Chloride(0) <= 103.10 AND Serum Chloride(0) <= 101.50 AND Ionized Calcium(0) > 1.15 | 210 | 0.26 | 0.97 | 6 | 7 | -1 |
| 12 | Serum Chloride(0) <= 103.10 AND Serum Chloride(0) > 101.50 | 74 | -0.03 | 0.86 | 13 | 8 | 5 |
| 13 | Serum Chloride(0) > 103.10 | 254 | 0.23 | 0.96 | 8 | 10 | -2 |
| 14 | Serum Albumin(1) > 3.15 AND Differences in Bicarbonate <= 3.15 AND Differences in Oxygen Saturation > -1.33 | 316 | -0.30 | 0.91 | 11 | 8 | 3 |
| 15 | Alkaline Phosphatase(1) <= 69.50 AND Serum ALT(1)<= 44 AND Age <= 69.60 | 110 | 0.20 | 0.98 | 3 | 18 | -16 |
| 16 | Hemoglobin(1) > 7.35 AND Ionized Calcium(1) <= 1.54 AND Differences in Lactate Dehydrogenase <= 24.92 | 611 | 0.69 | 0.96 | 9 | 12 | -3 |
| 17 | Serum ALT(1) <= 32 AND Differences in Serum Glucose <= -0.85 | 112 | 0.11 | 0.98 | 2 | 15 | -12 |
| 18 | Alkaline Phosphatase(0) > 76.50 AND Differences in Serum Glucose > -0.85 | 465 | -0.28 | 0.92 | 10 | 10 | 0 |

(0) Indicates value on the day of admission (1) Indicates value on the day prior to transplant Differences are defined between values on the day prior to transplant versus those on the day of admission

**Table 5.** RuleFit and Expert Rankings for the Secondary Outcome of Readmissions at 90-Days.

| Rule # | Rule Description | # Cases | Model Coefficient | Empirical Risk (Importance) | RuleFit Importance | Expert Importance | Rank Difference |
| --- | --- | --- | --- | --- | --- | --- | --- |
| 1 | Serum Phosphorus(0) > 3.25 AND Differences in Bicarbonate <= 2.41 AND Differences in Mean Arterial Pressures > -10.45 | 319 | -0.04 | 0.51 | 4 | 4 | 0 |
| 2 | Differences in Phosphorus > 8.50 | 42 | 0.21 | 0.76 | 1 | 2 | -1 |
| 3 | Blood Urea Nitrogen(0) > 22.50 AND Serum Glucose(1) <= 109.50 | 101 | 0.08 | 0.69 | 2 | 3 | -1 |
| 4 | Blood Urea Nitrogen(0) > 22.50 AND Serum Glucose(1) > 109.50 AND Respiratory Rate(0) <= 24.50 | 416 | -0.18 | 0.50 | 5 | 6 | -1 |
| 5 | Serum Calcium(1) > 9.05 AND Serum Magnesium(0) <= 2.05 AND Differences in Heart Rate <= 23.95 AND Differences in Platelet <= -7.50 | 164 | -0.36 | 0.42 | 6 | 4 | 2 |
| 6 | Serum Albumin(1) <= 4.65 AND Serum ALT(0) > 57.50 AND New Hemodialysis (Transplant versus Admission) | 117 | 0.12 | 0.68 | 3 | 1 | 2 |
| (0) Indicates value on the day of admission |  |  |  |  |  |  |  |
| (1) Indicates value on the day prior to transplant |  |  |  |  |  |  |  |
| Differences are defined between values on the day prior to transplant versus those on the day of admission |  |  |  |  |  |  |  |

### EAML Model Performance Versus Cross-Sectional and Other ML Models

For the primary outcome of death at one-year: AUROCs were 0.707 (Confidence Interval [CI] 0.625-0.793) for the EAML and 0.719 (CI 0.640-0.800) for the RuleFit models. For the secondary outcome of death at 90-days: AUROCs were 0.678 (CI 0.581-0.776) for the EAML and 0.707 (CI 0.615-0.800) for the RuleFit models.

Pairwise AUROC differences and confidence intervals are reported in detail in Figure 2 for the primary outcome of death at one-year and in Figure 3 for the secondary outcome of death at 90-days. In general, for the outcomes of death at one-year and death at 90-days, AUROC differences between EAML and RuleFit models showed that RuleFit outperformed EAML but this was not significant: Δ(RuleFit - EAML) was 0.013 (CI -0.027-0.052) for death at one-year and Δ(RuleFit - EAML) was 0.030 (CI -0.100- 0.071) for death at 90-days. Moreover, AUROC differences between the EAML/RuleFit models and GBM, and those between the EAML/RuleFit models and GLMNET were also not significant. In contrast, for the outcomes of death at one-year and death at 90-days, the EAML/RuleFit models consistently outperformed cross-sectional models (MELDNa, NACSELD, CLIF-ACLF, and TAM).

For the secondary outcome of readmission at 90-days: AUROCs were 0.557 (CI 0.493-0.623) for the EAML and 0.564 (CI 0.498-0.629) for the RuleFit models. Pairwise AUROC differences and confidence intervals are reported in detail in Figure 4 for the secondary outcome of readmission at 90-days. In general, the EAML and RuleFit models did not show significant differences in predictive abilities versus each other and versus other ML models. Moreover, while EAML/RuleFit showed significant differences in AUROC versus some of the cross-sectional models (MELDNa, NACSELD, and CLIF-ACLF) – overall predictive abilities of all models evaluated were poor.

**Figure 4.**
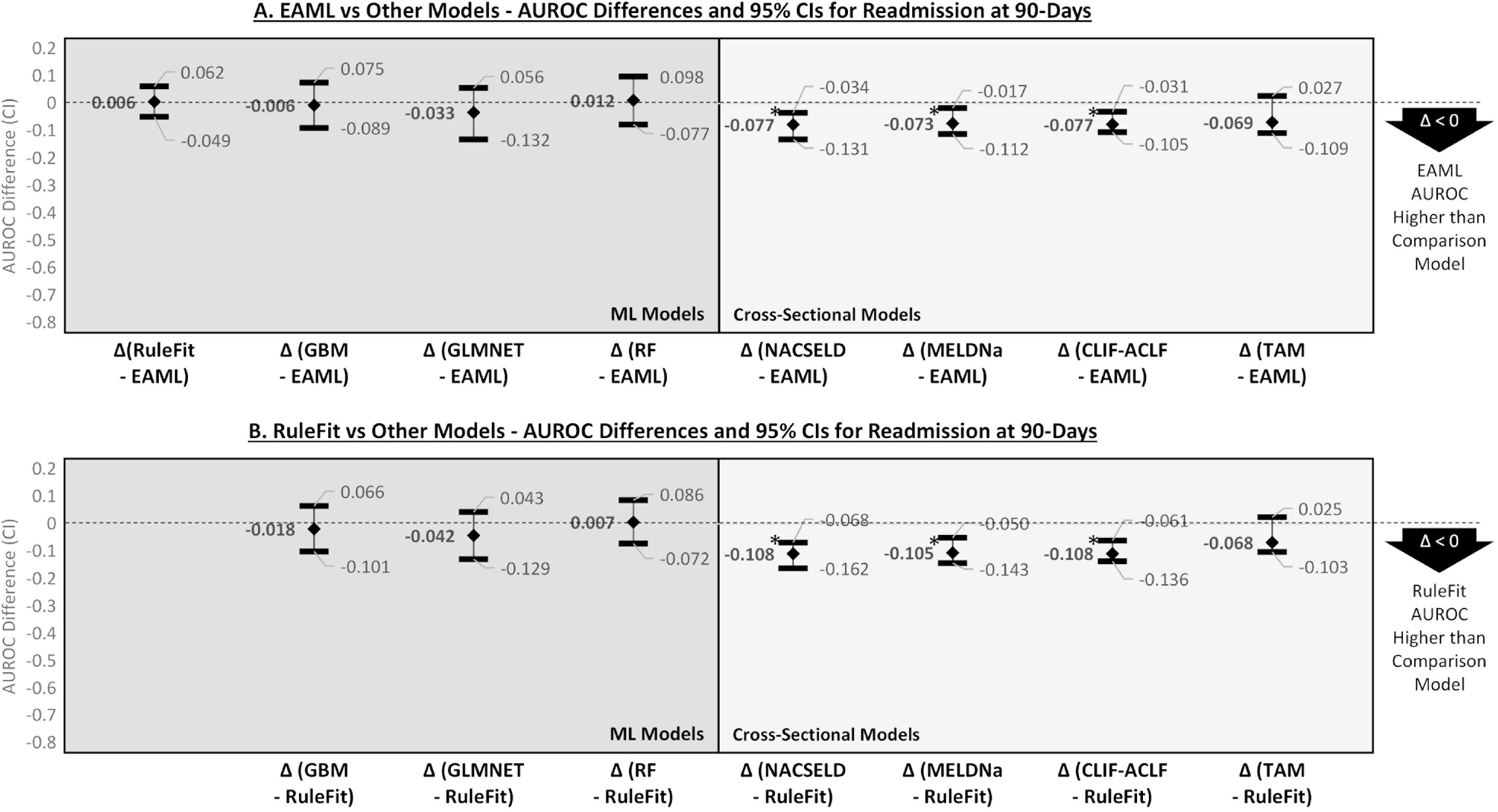
AUROC Differences and Confidence Intervals for EAML/RuleFit versus Other Models for the Outcome of Readmissions at 90-Days

## Discussion

This study is one of the first to explicitly combine human expert knowledge with ML to create an interpretable ML model for a clinical problem within transplantation. In this study, we generated two models (EAML, which incorporates human expert content, and RuleFit, which does not incorporate human input) for each of the three outcomes (post-transplant mortality at one-year, post-transplant mortality at 90-days, and readmission after transplant at 90-days). Our ML models (EAML and RuleFit) significantly outperformed existing cross-sectional models with mean AUROCs clustering around 0.700 for the outcomes of post-transplant mortality at one-year and mortality at 90-day.

In our pairwise comparisons of models utilizing AUROC differences, we found that while there were no significant differences between EAML and RuleFit, and between EAML/RuleFit and other popular ML algorithms, such as GBM and GLMNET. Moreover, while these were not statistically significant, but the EAML models consistently had lower AUROCs versus the RuleFit models. The most likely explanation in this situation is due to residual artifacts in study population as the training, validation, and test sets are all derived from the same database. In this circumstance, the process of incorporating expert input with EAML is not expected to improve the performance of the model since the test set have similar distributions of demographic and clinical characteristics as the training sets.

The purpose of EAML, therefore, in this situation is to reveal key insights from the discrepancies between human expert and ML rankings of rules. These reveal residual biases and areas for future research. For instance, in the EAML model for post-transplant mortality at one-year, rule #18 (MELDNa at the time of transplant being > 32.47) was ranked as the most important by experts, but only tenth most important by RuleFit. This difference in rank by nine positions indicated that experts may have biases favoring of a well-known and established clinical scoring system – whereas the RuleFit algorithm determined it to be not as important. In general, across the three outcomes, ACLF experts were more likely to over-rank the importance of commonly used physiologic and clinical makers, such as MELDNa, age, and white blood cell count. In contrast, RuleFit was more likely to elevate the importance of electrolytes and hematological parameters, such as ionized calcium, sodium, and lactate dehydrogenase as important data features. These results imply additional avenues for further research in the clinical care of patients with ACLF (Figure 5). Moreover, this study demonstrates that EAML’s use may not be limited to predictive modeling, but also as an artificial intelligence-guided method for hypothesis generation.

**Figure 5.**
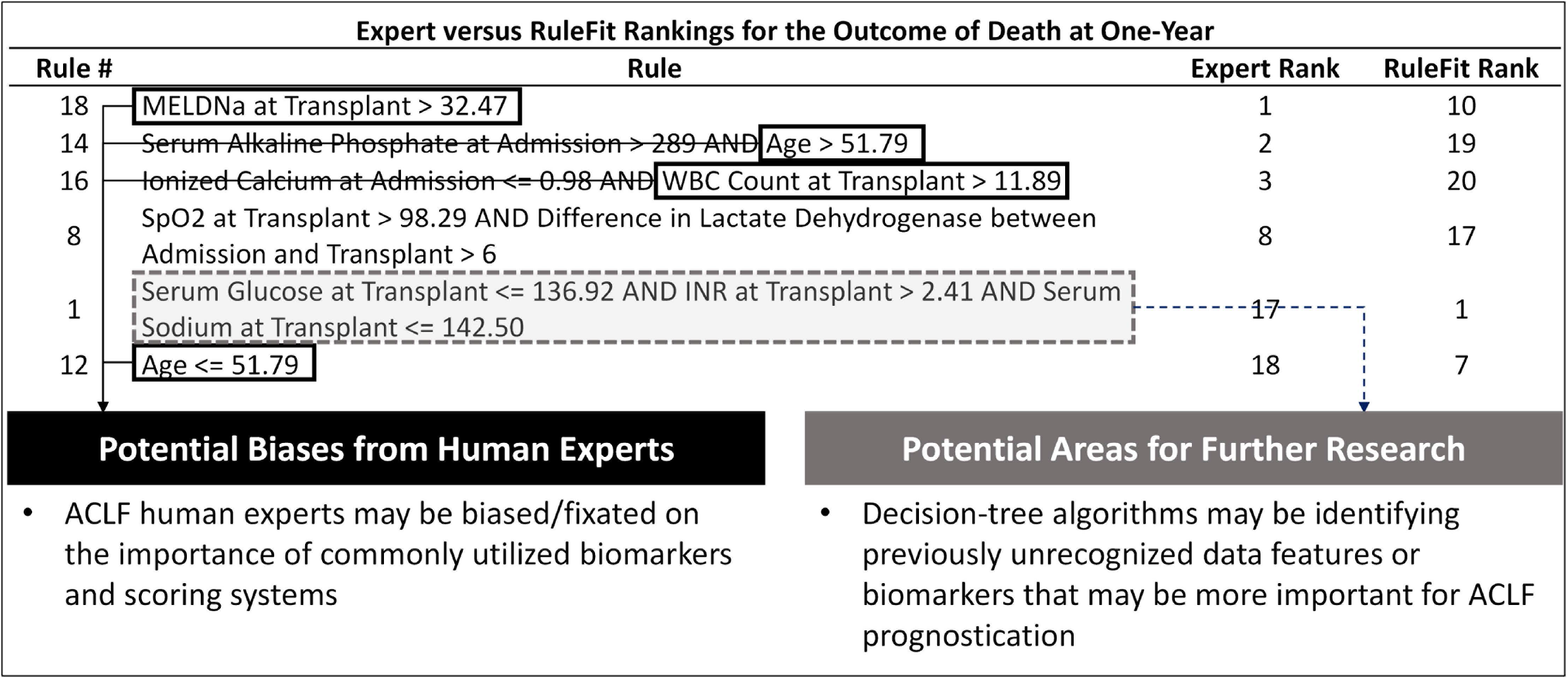
Disagreements Between Experts and RuleFit May Reflect Biases, Artifacts, and Areas for Further Research

Finally, this was the second study to fully utilize UCHDW, a novel big data multi-center EHR database, and the first to derive insights on transplant patients. UCHDW is based on the OMOP common data model, which is also utilized in several other big data multi-center EHR databases, such as the National COVID Cohort Collaborative (N3C),(44) All of Us,(45) and the Veterans Health Administration Corporate Data Warehouse (VHACDW).(46,47) While patients with ACLF and LT patients have been extensively studied in the VHACDW, the VHACDW is not broadly representative of the general population. While patients with cirrhosis have been studied in N3C, the current purviews of N3C limits research topics to those related to the novel coronavirus pandemic. It is our hope that our analytical approach of utilizing OMOP will become more common as increasing numbers of institutions have or are in the processing of harmonizing their EHR data to the OMOP common data model.

There are several limitations to this study due to its retrospective nature, its use of a novel database, and its analytical processes. First, there is selection bias – we had only included patients with ACLF who had successfully undergone LT, and not those who were listed to undergo LT but then subsequently died or recovered and not those who were never listed for LT. This means that the patients with ACLF who ultimately made it LT suffered from a survivorship bias and are unlikely to be representative of the entire ACLF population. While it is feasible to pull data for all patients with ACLF who did not undergo LT, we have no visibility into whether these patients were listed for LT and we would not be able to evaluate for the post-transplant outcomes of interest.

Second, we do not have intraoperative or donor derived data for the patients in our cohort. This, however, is not necessarily a significant limitation in our study as our intended goal was to derive a pre-LT model to predict post-LT outcomes. The ultimate clinical decision that this model would help with is to whether to proceed to LT for an ACLF patient. Third, to take advantage of the high-dimensional nature of UCHDW, we only sourced data from three transplant centers within UCH. In addition, all three UCH facilities included are in the state of California. While this population is demographically diverse, California has some of the highest MELDNa scores at the time of transplant. The models and their results, therefore, may not be generalizable to other settings. In addition, as ACLF etiologies may be variable across geographies, our models and conclusions may not be generalizable to populations outside the United States. These geographic-based differences may be a contributor to why the TAM model based on French ACLF patients performed poorly in our populations. External validation should be undertaken for these model prior to their potential deployment in clinical practice.

Finally, the analysis codes utilized to derive the data from UCHDW were written for this specific (UCHDW) implementation of the OMOP common data model. While OMOP is a common data model that allows for generalization of analyses across different datasets, there may be minor variations and differences in data structures, semantics, and coding. The OMOP-based extraction methods and algorithms for these analyses have not been tested on other OMOP-based data sources – further research is required to evaluate for true “out-of-the-box” interoperability.

Despite these limitations, this study represents “proof of concept” for several key conceptual developments for health services research in transplantation: 1. Use of human expert augmentation in ML modeling, 2. Generation of multiple ML models that outperforms traditional cross-sectional models for predicting post-transplant outcomes in ACLF, and 3. Utilizing of a novel data source and common data model in transplant hepatology. With further external validation, the EAML models generated in this study could be refined and evaluated in an iterative manner in clinical decision support (CDS) systems to actively guide clinical decision-making. In such a CDS-based implementation, prospective surveillance of outcomes would then allow for active feedback to further improve these models.

## Supporting information

Suppplemental Materials

## Data Availability

The authors thank the Center for Data-driven Insights and Innovation (CDI2) at the University of California Health (UCH) for its analytical and technical support related to the use of the UC Health Data Warehouse (UCHDW) and related data assets, including the UC COVID Research Data Set (CORDS).

## Abbreviations

ACLF: acute-on-chronic liver failure;
APASL ACLF: Asian Pacific Association for the Study of the Liver ACLF Research Consortium;
AUROC: area-under-the-receiver-operating characteristic curve;
CDI2: Center for Data-Driven Insights and Innovation;
CDS: clinical decision support;
CI: confidence interval;
CORDS: UC COVID Research Data Set;
CPT4: Current Procedural Terminology version 4;
EAML: Expert-Augmented Machine Learning;
EF-CLIF: European Association for the Study of the Liver-Chronic Liver Failure Consortium;
EHR: electronic health record;
ESLD: end-stage liver disease;
FiO2: fraction of inspired oxygen;
GBM: Gradient Boosting Machine;
GLMNET: Elastic-Net Regularized Generalized Linear Model;
ICD-10-CM: International Classification of Diseases, Tenth Revision, Clinical Modification;
LASSO: Least Absolute Shrinkage and Selection Operator;
LT: liver transplantation;
ML: machine learning;
N3C: National COVID Cohort Collaborative;
NACSELD: North American Consortium for the Study of End-Stage Liver Disease;
OMOP: Observational Medical Outcomes Partnership;
PaO2: arterial partial pressure of oxygen;
RF: Random Forest;
SNOMED: Standard Nomenclature of Medicine;
SpO2: partial oxygen saturation;
TAM: Transplantation for ACLF-3 Model;
UCH: University of California Health;
UCHDW: University of California Health Data Warehouse;
VHACDW: Veterans Health Administration Corporate Data Warehouse

## Data Acknowledgement

The authors thank the Center for Data-driven Insights and Innovation (CDI2) at the University of California Health (UCH),(1) for its analytical and technical support related to the use of the UC Health Data Warehouse (UCHDW) and related data assets, including the UC COVID Research Data Set (CORDS).

## Notes

**Financial/Grant Support:** The authors of this study were supported in part by the KL2TR001870 (National Center for Advancing Translational Sciences, Ge and McCulloch), AASLD Anna S. Lok Advanced/Transplant Hepatology Award AHL21-104606 (AASLD Foundation, Ge), American Society of Transplantation LICOP Grant Award CA-0182782 (American Society of Transplantation, Ge), P30DK026743 (UCSF Liver Center Grant, Ge and Lai), F31HL156498 (National Heart, Lung, and Blood Institute, Digitale), UL1TR001872 (National Center for Advancing Translational Sciences, Pletcher), and R01AG059183/K24AG080021 (National Institute on Aging, Lai). The content is solely the responsibility of the authors and does not necessarily represent the official views of the National Institutes of Health or any other funding agencies. The funding agencies played no role in the analysis of the data or the preparation of this manuscript.

**Disclosures:** Dr. Jin Ge receives research support from Merck and Co.

### Competing Interest Statement

Dr. Jin Ge receives research support from Merck and Co.

### Funding Statement

The authors of this study were supported in part by the KL2TR001870 (National Center for Advancing Translational Sciences, Ge and McCulloch), AASLD Anna S. Lok Advanced/Transplant Hepatology Award AHL21-104606 (AASLD Foundation, Ge), American Society of Transplantation LICOP Grant Award CA-0182782 (American Society of Transplantation, Ge), P30DK026743 (UCSF Liver Center Grant, Ge and Lai), F31HL156498 (National Heart, Lung, and Blood Institute, Digitale), UL1TR001872 (National Center for Advancing Translational Sciences, Pletcher), and R01AG059183/K24AG080021 (National Institute on Aging, Lai). The content is solely the responsibility of the authors and does not necessarily represent the official views of the National Institutes of Health or any other funding agencies. The funding agencies played no role in the analysis of the data or the preparation of this manuscript.

### Author Declarations

The Institutional Review Board at the University of California, San Francisco gave ethical approval for this work under #20-32717 for model generation and #22-37555 for expert input.

