## Supplementary material for "Predicting Post-Liver Transplant Outcomes in Patients with Acute-on-Chronic Liver Failure using Expert-Augmented Machine Learning": Suppplemental Materials

**Supplemental Methods:**

*The Observational Medical Outcomes Partnership (OMOP) Data Model*

OMOP is an open-community and common data model to enable standard analyses of observational databases. In the OMOP common data model, classification vocabularies, such as International Classification of Diseases, Tenth Revision, Clinical Modification (ICD-10-CM), Current Procedural Terminology version 4 (CPT4), or Standard Nomenclature of Medicine (SNOMED); are mapped to standard OMOP concepts based on semantic and clinical relationships.(1) Vocabulary classification and mapping of various ontologies to the OMOP standard vocabulary is maintained by OHDSI and publicly available on ATHENA (http://athena.ohdsi.org/), which is a web-based vocabulary repository.(2)

*Definitions of ACLF*

The NACSELD ACLF diagnostic criterion is based on two or more organ failures: shock, West-Haven grade III/IV hepatic encephalopathy, dialysis, and/or mechanical ventilation.(3) The EF-CLIF ACLF diagnostic criterion is based on gradations of organ failures based on laboratory measurements and/or events across six organ systems.(4) As the ratio of arterial partial pressure of oxygen (PaO2) to fraction of inspired oxygen (FiO2) was not always available, we utilized the threshold of partial oxygen saturation (SpO2) to FiO2 ratio of ≤ 214 as an equivalent of PaO2/FiO2 ratio of ≤ 200 as per custom in the CLIF-C-ACLF model.(4)

*Missingness and Imputation*

Implausible values, as defined as those greater than three standard deviations from the mean values were removed as per previous data processing in the generation of ML models.(5) Data features and variables with greater than or equal to 25% missingness were excluded from analyses, similar to previous ML analyses of transplant hepatology patients.(5) Data features with less than 25% missingness were imputed with single imputation with chained random forests as implemented in the *missRanger*, version 2.1.3, R package.(6,7)

This imputation method handles both continuous and categorical data features within the distribution of the original data, thereby avoiding outliers and recovers the natural data variability. In previous performance evaluations of imputation methods in a large electronic health record dataset, *missRanger* algorithms produced the most reliable results with the lowest average standard errors. (8–10)

*Confidence Interval Calculations for AUROC Differences*

To estimate point estimates and confidence intervals for AUROC differences between pair-wise comparisons of models (e.g. EAML versus MELDNa), we utilized bootstrapping over 2,000 iterations for each difference as implemented in the *boot*, version 1.3-28, R package.(11–13)

**Supplemental Figure 1 – RuleFit Training and Testing Plot for Outcome of Death at One-Year**

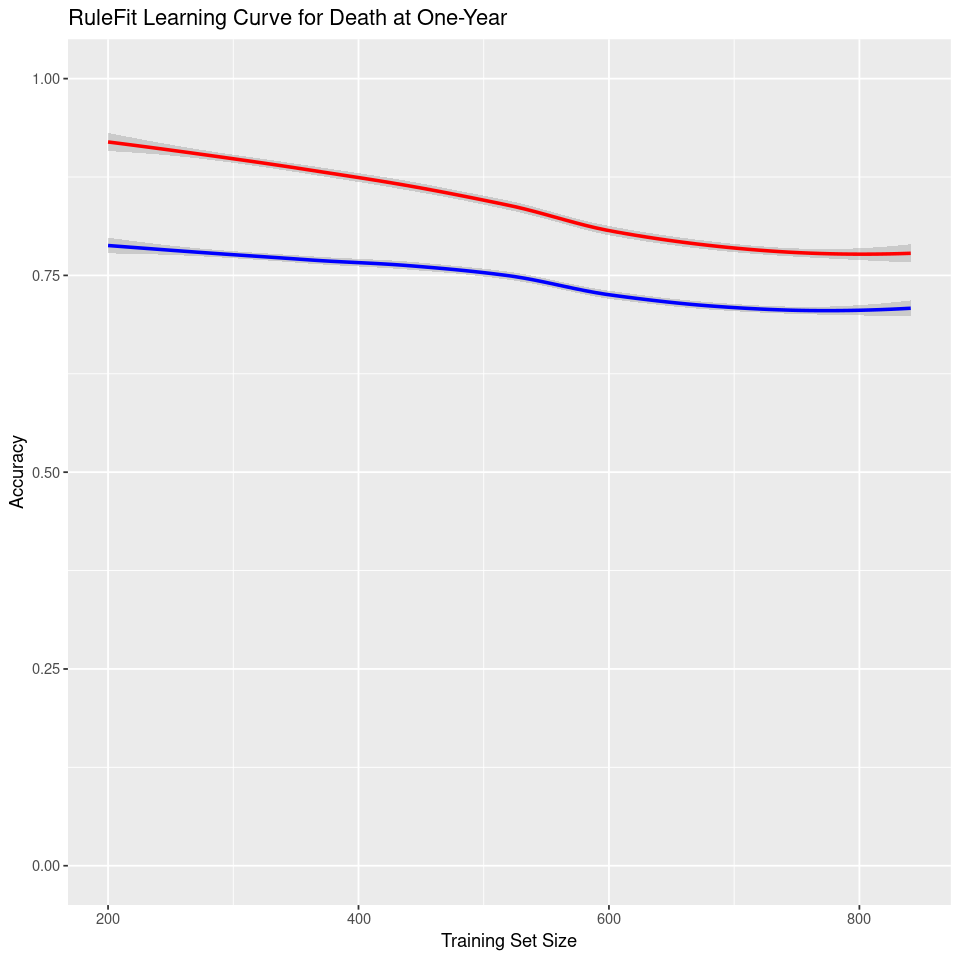

Training Plot

Test Plot

**Supplemental Figure 2 – RuleFit Training and Testing Plot for Outcome of Death at 90-Days**

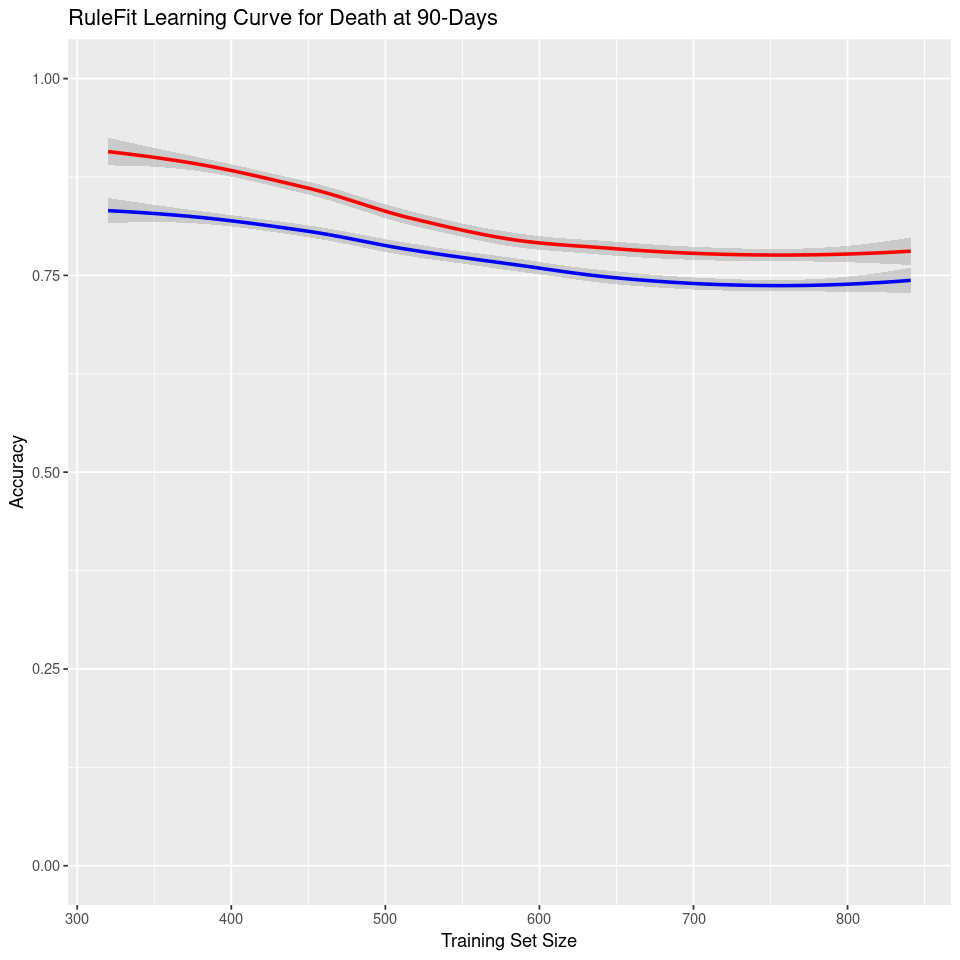

Training Plot

Test Plot

**Supplemental Figure 3 – RuleFit Training and Testing Plot for Outcome of Readmissions at 90-Days**

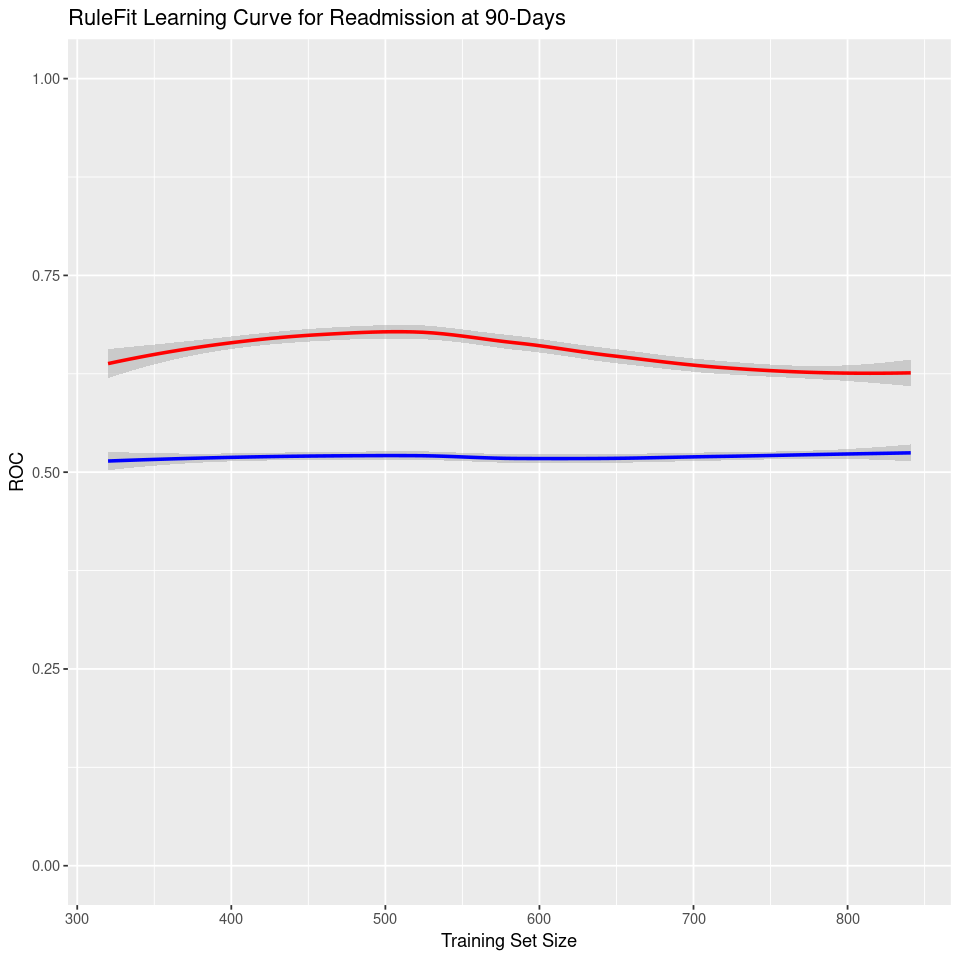

Training Plot

Test Plot

**Supplemental Materials References:**

10.	Liu D, Oberman HI, Muñoz J, Hoogland J, Debray TPA. Quality control, data cleaning, imputation. arXiv. 2021;

11.	Bootstrap Functions (Originally by Angelo Canty for S) [R package boot version 1.3-28.1] [Internet]. 2022 [cited 2023 Jan 6];Available from: https://cran.r-project.org/web/packages/boot/index.html

12.	Carpenter J, Bithell J. Bootstrap confidence intervals: when, which, what? A practical guide for medical statisticians. Stat. Med. 2000;

13.	DiCiccio TJ, Efron B, Hall P, Martin MA, Canty AJ, Davison AC, et al. Bootstrap confidence intervalsCommentCommentCommentCommentRejoinder. Stat Sci. 1996;11:189–228.
